# The Bupa Arabia Value Pilot: A KSA-Based Payor-Provider Collaborative in Maternity Care

**DOI:** 10.1101/2025.06.07.25328955

**Authors:** Abdulrehman Taher, Serena Sarkis, Hussein H. Khachfe, Fares Saade, Jason Greich, Eman ElBadrawy, Dalal AlHamed

**Author notes:** <u>Corresponding Author:</u> Hussein H. Khachfe, MD, PMP, Capadev LLP, London, United Kingdom. **Authorship Confirmation:** All authors certify that they meet the criteria for authorship.

## Abstract

The Bupa Arabia and Bupa CareConnect Value Pilot explores the transformative journey of implementing Value-Based Healthcare (VBHC) in Maternity care across four hospitals in Saudi Arabia, using ICHOM’s Pregnancy and Childbirth Standard Set. Led by Bupa Arabia, a leading insurance provider in the Kingdom, with implementation and technology support from Capadev, an ICHOM implementation and accreditation partner, this initiative showcases the power of a multi-stakeholder collaboration in improving outcomes for expectant mothers. The paper offers insight into the processes, digital tools, and strategies for engaging patients and clinicians, and highlights both the challenges and lessons learned during the first nine months of implementation, while sharing early results aimed at improving care quality.

## Introduction

Aligned with Saudi Arabia’s Vision 2030 and its commitment to healthcare transformation around Value-Based Healthcare (“VBHC”) principles, Bupa Arabia, the leading health insurer in the Middle East, is leading the way by implementing VBHC and driving towards sustainability in care quality and efficiency. This includes strengthening capabilities in outcomes measurement and exploring innovative payment models (Chowdhury S, 2021). As part of this strategic shift, Bupa Arabia launched a groundbreaking payer-provider collaboration pilot with four leading tertiary care providers, focusing on Maternity care and adhering to the International Consortium for Health Outcomes Measurement (“ICHOM”) standard sets.

This pilot, conducted in partnership with Capadev, a licensed ICHOM implementation and accreditation partner (Capadev Homepage, n.d.), was designed to achieve the following goals:

1. Standardize care pathways and outcome measurement to enable accurate benchmarking.
2. Implement the collection of Patient-Reported Outcome Measures (PROMs) alongside Clinician-Reported Outcome Measures (CROMs), based on the ICHOM standard sets.
3. Translate data into actionable insights to drive inform clinical decision-making, improve care quality and efficiency, and address female varying needs in Maternity care.
4. Integrate patient-centered outcomes as a key component of the framework used to identify tertiary providers as Centers of Excellence.
5. Apply lessons learned to scale VBHC initiatives and contribute to the broader VBHC transformation in the Kingdom.
6. Explore opportunities for innovative payment models, including value-based bundled payment approaches.

Maternity was selected as the primary focus for the first VBHC pilot based on multiple criteria, including:

1. Alignment with the National PROMs (NPROMs) strategy outlined by the Council for Health Insurance (CHI) (Council for Health Insurance, 2021).
2. Availability of well-defined ICHOM standard sets (ICHOM, n.d.).
3. High volume combined with wide variations observed, implying significant impact potential of a VBHC approach.

## Methodology

The pilot was structured into five phases:

### Pre-Launch Planning

□ Four providers were selected based on volume, leadership and clinical commitment, and technology readiness
□ Site Rollout Teams (SRTs), comprising physicians, nurses, and quality representatives, were formed to lead the initiative at each provider site.
□ SRTs participated in a blended online and in-person course on VBHC to build their capabilities and knowledge. Members received a certificate after successfully completing the course.
□ SRTs developed Patient Journey Maps optimized based on global best practices, which guided the definition of outcomes collection points.

### Outcomes Rollout Preparation

□ Ten PROMs instruments from ICHOM standard sets were selected to track maternal health, covering general health (EQ-5D-3L), sexual functioning (PROMIS SexFS), mental state (PHQ-2, EPDS), urinary incontinence (ICIQ), stool incontinence (Wexner), birth satisfaction (BSS-R), mother-infant bonding (MIBS), breastfeeding (BSES), social support (SIMSS), and postpartum depression (EPDS) as shown in Figure 1.
□ Tools were localized following ISPOR guidelines and validated for Saudi cultural relevance through Acolad, an approved ICHOM partner.
□ CROMs were also selected as part of ICHOM standard sets.
□ Case-mix variables were selected based on the ICHOM Pregnancy and Childbirth Standard Set to ensure consistency with global best practices.
□ Five key control points were defined to guide the collection schedule: first visit, third trimester, immediate post-delivery, two weeks post-delivery, and six months post-delivery.
□ Patient inclusion criteria were established to ensure comparability across providers: women aged 18–45 years, Arabic speakers, residents of specified regions, no significant comorbidities, less than 24 weeks pregnant at first data collection, and no prior health complications.
□ This study was reviewed and approved by the Bupa Arabia Ethics, Value Based Healthcare and Cyber Security Committees, affiliated with Bupa Arabia for Cooperative Insurance Company, Jeddah, Saudi Arabia. The committee evaluated the full study protocol, including data collection procedures for Patient-Reported Outcome Measures (PROMs), Clinician-Reported Outcome Measures (CROMs) and consent forms shared across participating sites. The committee issued approval based on the anonymized and aggregated nature of the data collected, the use of secure digital platforms, and adherence to local data privacy regulations in Saudi Arabia.

**Figure 1.**
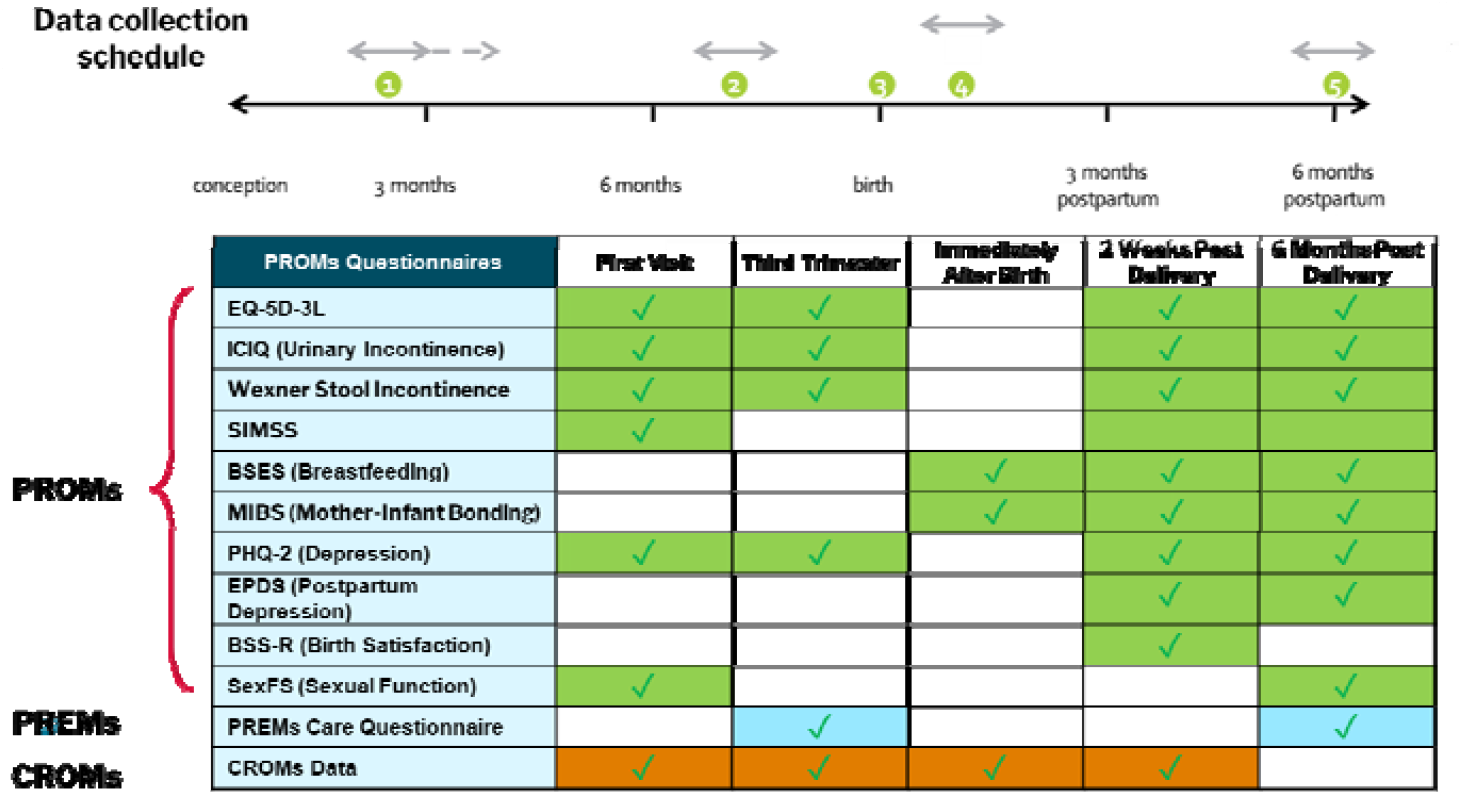
Outcomes to be collected and control point schedule

### Data Collection Solution Launch

□ Capadev’s platform was configured and deployed on a Level C KSA-compliant local server.
□ The platform was implemented across all four providers to enable digital, automated outcomes collection.
□ PROMs assessments were automatically sent to females via their preferred communication method (SMS, WhatsApp, email).
□ Each assessment included a consent step to ensure female approval and data confidentiality.
□ Clinicians used standardized Excel templates to input and upload CROMs.
□ The platform aggregated data, enabling real-time analysis, dashboarding, and provider/physician-level reporting.

### Outcomes Rollout

□ A female communication and onboarding plan was co-developed with stakeholders to enhance engagement and response rates.
□ PROMs were collected, and CROMs were uploaded to the platform in accordance with ICHOM standards.
□ Continuous expert monitoring addressed site-specific challenges, ensured compliance with data collection accuracy, and maintained high data quality at the source.

### Dashboarding and Ramp-up

□ Data was analyzed to develop dashboards and identify actionable value-driven insights aimed at improving outcomes and reducing costs.
□ SRTs were trained to derive actionable insights from dashboards and systematically use outcomes to optimize care quality and its patient-centricity (Figure 2).
□ After the first year, a data review will be conducted to assess completeness, validity, and accuracy, with accreditation granted to sites meeting the criteria.

**Figure 2.**
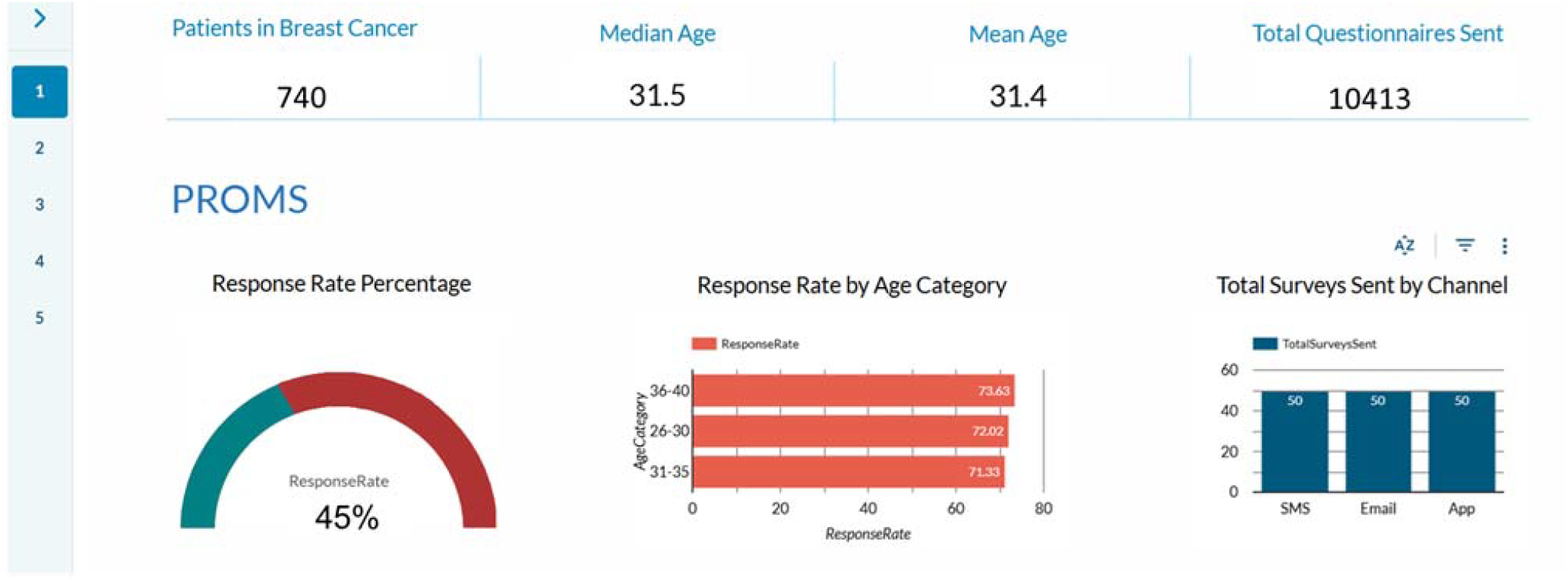
Digital platform dashboards (Illustrative)

## Results and Discussion

The outcomes rollout was launched in July 2024. Over the course of nine months, a total of 740 females were onboarded, with 45% completing questionnaires and 36% having CROMs uploaded on their behalf (Figure 3).

**Figure 3.**
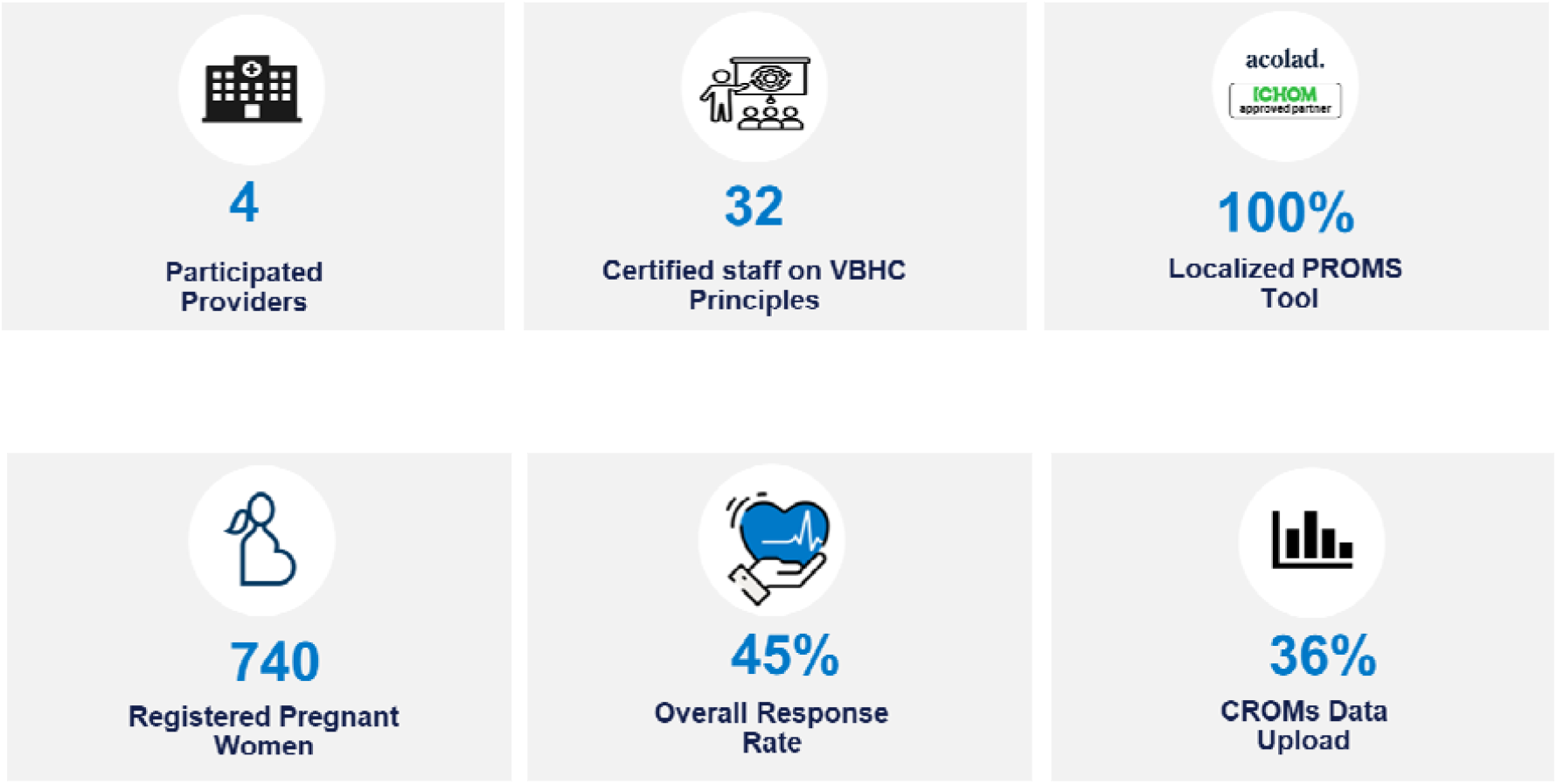
Milestones of Maternity Pilot

### Pre-Launch Planning Phase Observations

#### Variation in Maternity Care Delivery Practices Across Providers

An analysis of maternity care across four providers revealed one consistent practice regarding pregnancy confirmation:

□ Pregnancy Confirmation: All providers adhere to Ministry of Health (MoH) guidelines, offering an ultrasound to any woman with a positive HCG test to assess for a viable intrauterine pregnancy and to confirm or adjust the estimated date of delivery based on the last menstrual period. Pregnancies were presumed intrauterine unless the woman presented with one or more of the following risk factors for ectopic pregnancy: adnexal pain, vaginal spotting/bleeding, or other clinical indicators (Vadakekut & Gnugnoli, 2025).

However, three key variations were observed along the maternity care journey:

□ Prenatal Screening: Approaches to Non-Invasive Prenatal Testing (NIPT) varied. Some providers offered NIPT universally, while others applied a selective, risk-based approach. A risk-based model can help optimize resource use, but there is a need to harmonize guidelines for ultrasound and microbiological testing to ensure alignment with evidence-based practice.
□ Third-Trimester Ultrasound: There was inconsistency in the number of ultrasounds conducted in the third trimester. Two providers performed two scans, whereas the other two followed the standard practice of a single scan.

Addressing these variations will help improve maternal and neonatal outcomes while fostering a more consistent, value-based approach to maternity care.

#### Different Levels of Understanding of VBHC and PROMs

Providers exhibited varying levels of awareness and understanding of Value-Based Healthcare (VBHC) principles and the benefits of collecting PROMs. To ensure alignment, the structured learning journey—including e-learning and live training—ensured harmonization to build a common foundation and readiness for outcome measurement. Without such alignment, there is a risk that providers interpret and use PROMs inconsistently, leading to fragmented patient care, suboptimal care planning, and missed opportunities for continuous quality improvement based on holistic outcome trends. Without such alignment, several key risks can arise. For example:

□ Low Data Quality or Engagement: If providers don’t see the value, they may not encourage females to participate meaningfully leading to incomplete or poor-quality data.
□ Fragmented patient care, suboptimal care planning, and missed opportunities for continuous quality improvement based on holistic outcome trends.

#### Limited Utilization of Digital Channels in Maternity Care

The pilot revealed a lack of digital tools in maternity care delivery among some providers compared to others within the pilot, affecting integrated care, monitoring, and overall female experience. Strengthening digital adoption could enhance care coordination and female guidance throughout the maternity journey, while ensuring sustainable collection and usage of outcomes beyond this pilot.

### Outcomes Rollout Phase Observations

#### Response Rates

##### Per Control Point

A total of 740 females were registered on the platform, with an average response rate of 45%. This rate aligns with global benchmarks of PROMs data collection, which range between 30% and 50% (Figure 4).

**Figure 4.**
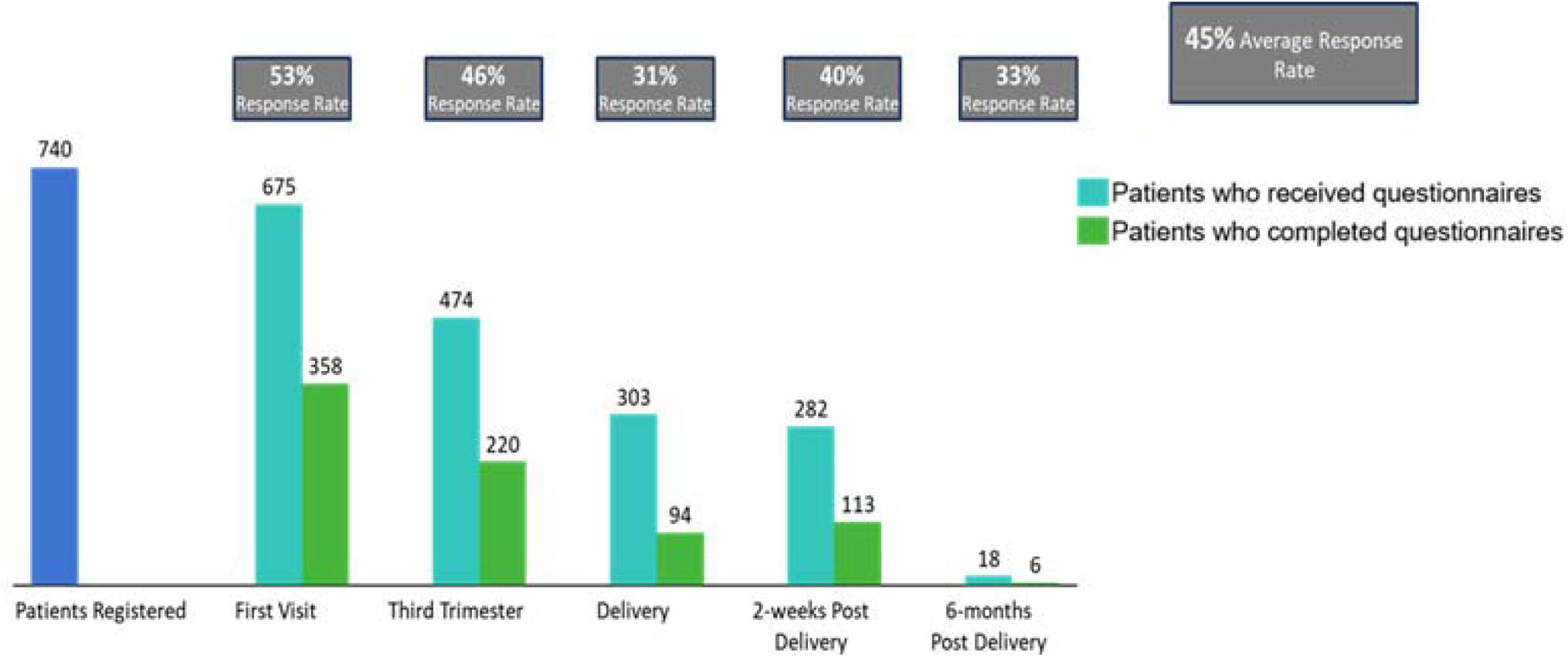
PROMs Response Rate

Response rates tend to decline as females progress through their journey, which can be attributed to several factors:

□ Inadequate education during the pilot onboarding process, where females may not fully understand the importance of responding to all control points throughout the journey.
□ Insufficient follow-up with patients to encourage continued engagement.
□ Fatigue and loss of interest, particularly after the third control point (immediately post-delivery), as females are preoccupied with other matters.
□ Suboptimal physician engagement in the process, particularly in making explicit use of the PROM observations.
□ Underutilization of collected data throughout the patient journey—particularly within clinical settings by the care team—to enhance female care.

##### Per Questionnaire

Analysis of response rates across different PROMs categories revealed varying levels of participation (Figure 5).

**Figure 5.**
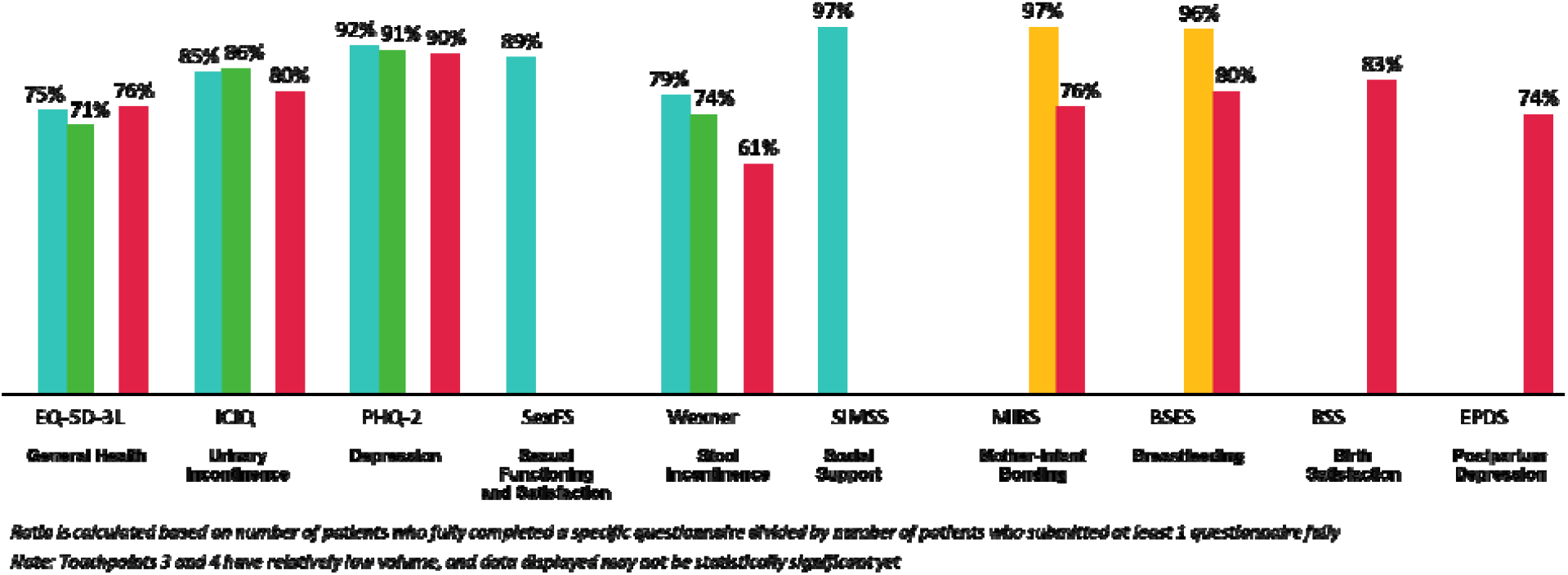
Average Individual PROMs Response Rate

To illustrate, PROMs related to Depression, Sexual Functioning and Satisfaction, and Social Support have the highest response rates, exceeding 89%.

In contrast, PROMs for General Health and Stool Incontinence record the lowest response rates, dropping to a low of 71% in some instances.

Contrary to the initial hypothesis that females would avoid sensitive topics like depression and sexual satisfaction, these domains achieved higher response rates. Women appear to feel more comfortable vocalizing such issues through a private and structured questionnaire rather than addressing them verbally during clinical visits. This reflects PROMs’ critical role in enabling pregnant women to raise concerns they may otherwise hesitate to express.

##### Per Hospital

Response rates among providers in the pilot program have ranged from 33% (within the global benchmark) to 66% (above the global benchmark), as shown in the chart below (Figure 6).

**Figure 6.**
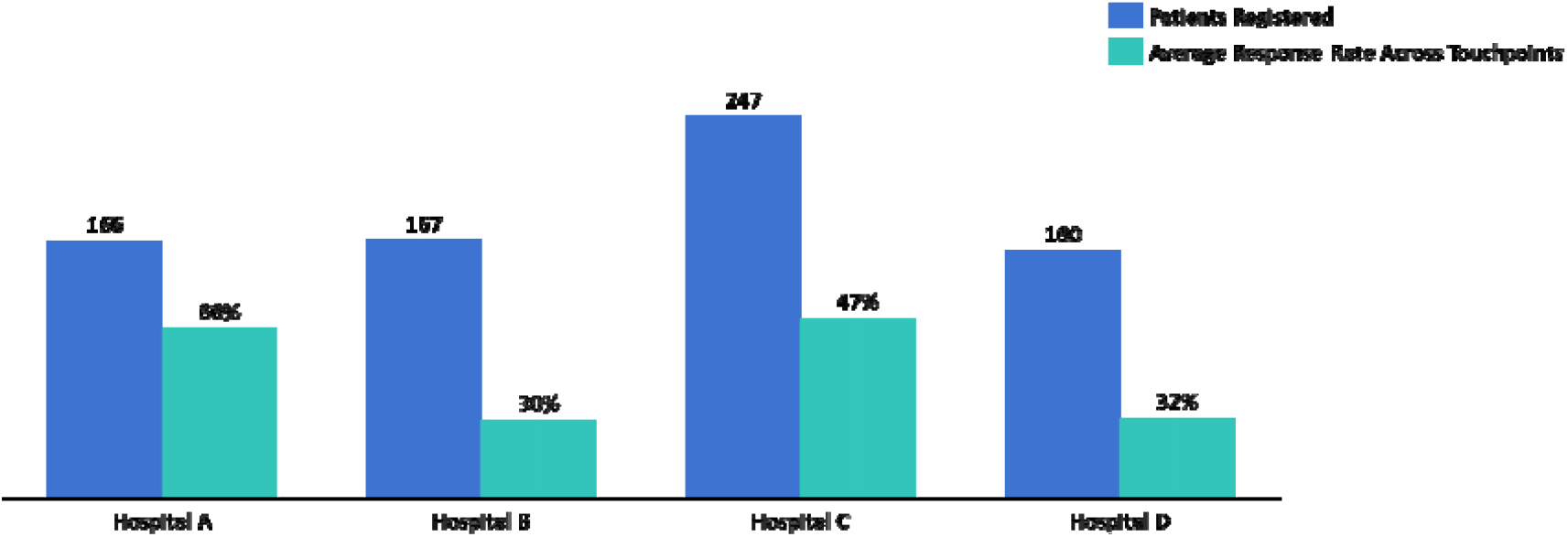
Average Response Rate per Hospital

Five key levers (Figure 7) were leveraged by providers to achieve response rate targets. It is evident that providers with the most levers implemented yielded the highest response rates. To illustrate, Provider A achieved the highest response rate (66%) by utilizing four of the five levers, including a structured onboarding process, consistent female engagement, and strong leadership support from the chief of the OBGYN department, who actively engaged the broader clinical team. Provider C followed with a 47% response rate, driven by initiatives such as a hospital-wide town hall to raise PROMs awareness, physician engagement, and consistent use of collected data to inform care improvements. Provider D reached a 32% response rate, supported by a dedicated VBHC team but hindered by irregular follow-up and limited use of collected data. Provider B recorded the lowest response rate (30%), relying mainly on an Integrated Practice Unit (IPU) that incentivized participation through benefits such as a private entrance and reduced waiting times contingent on PROMs completion, but lacked broader clinical team involvement.

**Figure 7.**
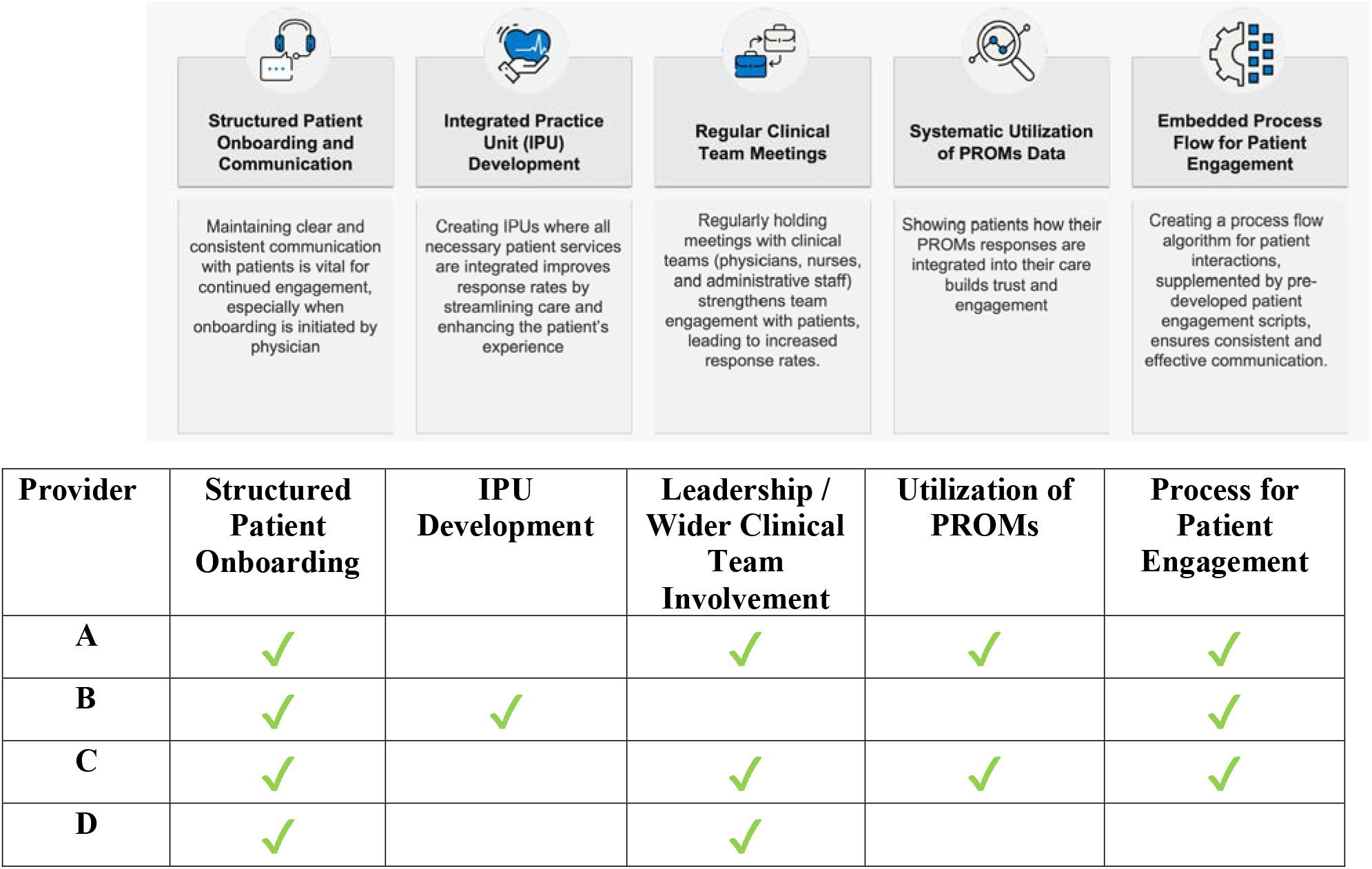
Key Levers Used to Improve Response Rate

These findings highlight that a multi-lever approach, especially leadership involvement and continuous use of data, is critical to maximizing female engagement and response rates.

## Data Analysis

To support clinical action and continuous quality improvement, all Site Rollout Teams (SRTs) were given access to real-time dashboards through the platform, displaying aggregated PROMs data. These dashboards used a color-coded system—green indicating normal or expected responses requiring no intervention, and red highlighting abnormal responses that warranted clinical follow-up. By visually flagging areas of concern such as elevated depression scores or low breastfeeding confidence, the dashboards enabled SRTs to promptly identify at-risk females, adjust care plans, and engage relevant clinical teams. This real-time visibility empowered providers to transition from passive data collection to active, outcomes-driven maternity care. A sample view of the dashboard is shown in Figure 8.

**Figure 8.**
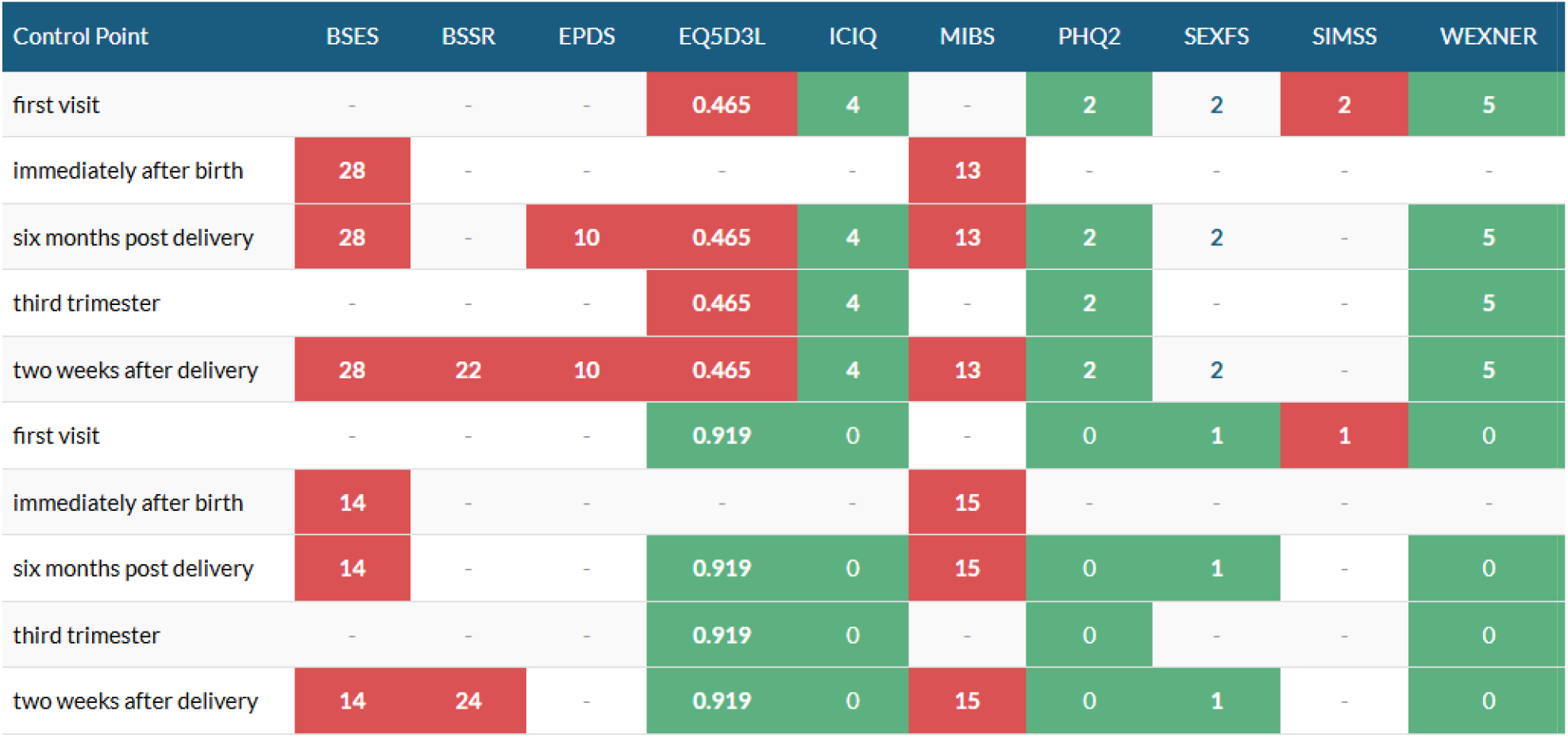
Score Analysis on Digital Platform Dashboard

### Overall Insights on Abnormal Responses

Abnormal responses were identified, most prominently in the following questionnaires (Figure 9):

**Figure 9.**
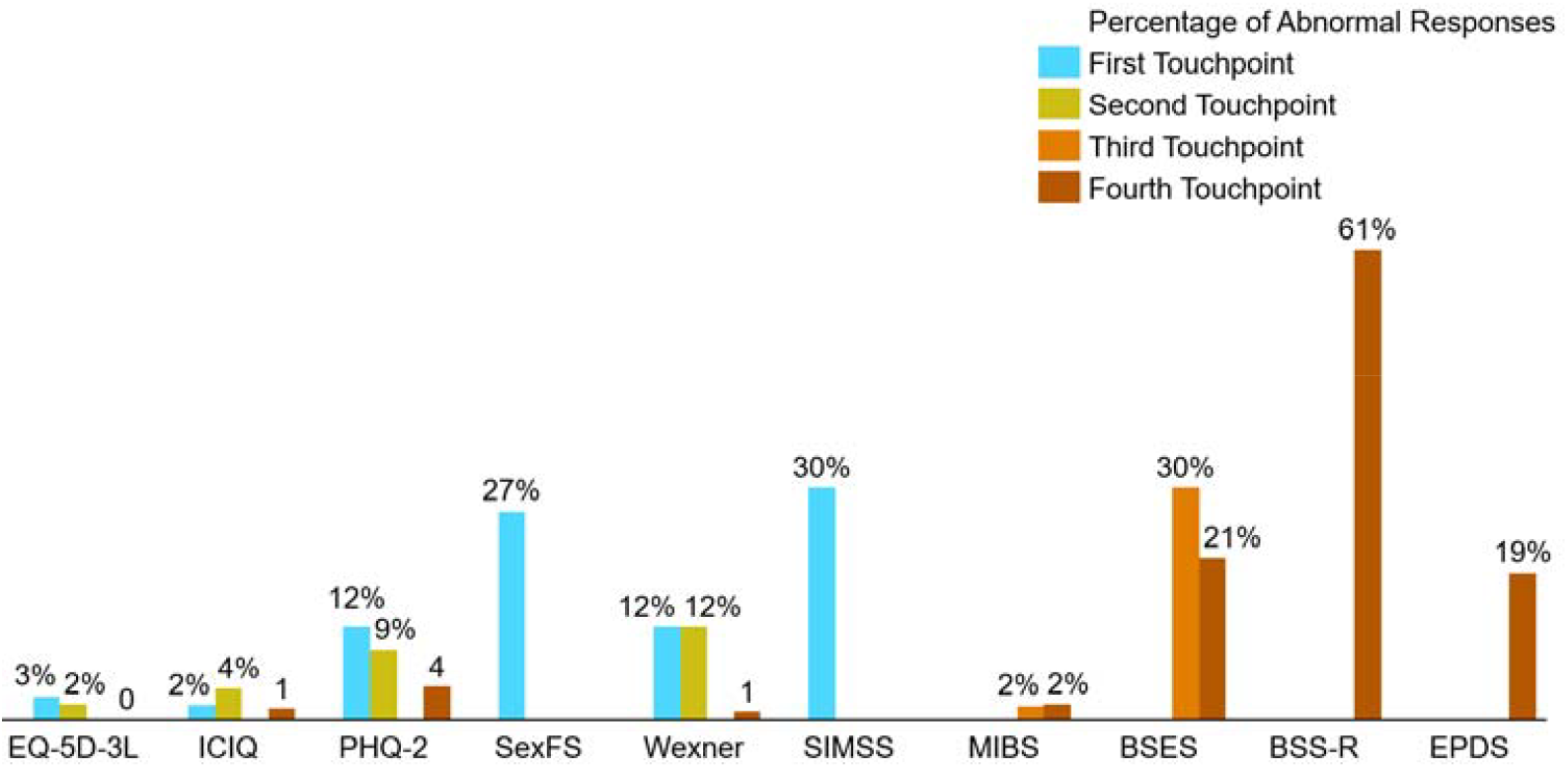
Abnormal Responses Among Questionnaires Across Control Points

#### First two control points

□ Sexual Functioning and Satisfaction: Reduced sexual satisfaction was reported by 27%, making it the second most common concern reported by pregnant women in the pilot.
□ Depression: A notable proportion of women (approximately 12% at the initial assessment) reported symptoms of depression or anxiety early in their pregnancies. This was followed by 19% of women expressing concerns consistent with postpartum depression.
□ Incontinence: Around 12% of women reported stool incontinence along the antenatal part of their pregnancy.

#### Last three control points

□ Birth satisfaction: The most common issue reported among postpartum women was a lack of birth satisfaction, with an overwhelming majority (61%) reporting this issue. Further investigation into the PROM answers themselves revealed that most females reported lack of control during the birthing process and excessive length of birth process to be the main issues affecting their experience.
□ Breastfeeding: Almost 30% of women reported issues related to breastfeeding in the immediate postpartum period.

##### PROMs Improvement Over Time: Control Points and Period Trends

PROMs were analyzed both across control points (CP1 to CP4) and between two phases of implementation: the **Early Period (June–October)** and the **Late Period (November–March)**. For the former, average responses per questionnaire per control point were calculated for all females and compared across control points—specifically for those measures repeated at different time points. For the latter, the average PROMs questionnaire responses across all control points for all females were compared between the first and second halves of the project, based on the rationale that providers implemented various initiatives to improve overall scores. The following section presents a combined view of the findings by questionnaire, highlighting how symptom trajectories evolved over time and across project phases.

#### EQ-5D-3L (General Health-Related Quality of Life)

EQ-5D-3L scores remained relatively stable across both control points and time periods. Between CP1 and CP2, average scores held steady (0.77 → 0.78), and similarly, from the Early to Late periods, scores shifted minimally from 0.76 to 0.78. These findings suggest that while individual symptoms (e.g., depression or incontinence) may fluctuate, women generally maintained a consistent perception of their overall health status. This also underscores the importance of interpreting condition-specific PROMs alongside general well-being measures to get a complete picture of female experience.

#### ICIQ-UI (Urinary Incontinence)

Urinary symptoms showed limited improvement across both analyses. From CP1 to CP2, scores remained virtually unchanged (3.70 → 3.71), indicating persistent symptoms during early recovery. However, comparing the Early to Late periods revealed a slight improvement (4.08 → 3.62), suggesting better symptom management across providers over time. These mixed results may reflect the while symptoms are not improving per female at provider sites from the first to second control point, proper screening and subsequent management with interventions can generally improve baseline urinary incontinence status among pregnant females, as other studies have shown (Alagirisamy P, 2022). One of the providers in the pilot introduced a referral pathway to a specialized pelvic floor rehabilitation center within their system for female patients who reported abnormal scores on the ICIQ-UI PROMs assessments. As a result, their average ICIQ-UI scores improved significantly—from 2.02 at the first touchpoint to 0.8 by the fourth. In contrast, other centers that did not implement similar interventions showed less improvement.

#### PHQ-2 and EPDS (Mental Health)

Mental health indicators consistently improved across both control points and phases. PHQ-2 scores declined steadily from CP1 to CP4 (1.18 → 0.75), indicating progressive emotional recovery. This trend was mirrored over time, with PHQ-2 and EPDS scores showing a downward shift between the Early and Late periods. These improvements likely reflect the impact of targeted psychiatric referral protocols introduced by some provider sites following early insights from PROMs data. This demonstrates how real-time feedback loops can guide timely interventions and improve emotional outcomes, which can improve maternal health in antenatal and postpartum phases (Chauhan A, 2022).

#### SexFS (Sexual Function and Satisfaction)

Sexual health scores remained largely stable across both analyses. Between the Early and Late periods, average scores shifted only slightly (2.11 → 2.03). This suggests that sexual function may be more influenced by underlying personal, relational, or physical factors at presentation than by short-term clinical interventions during the antenatal period. This corroborates observations from other authors on the complex factors affecting sexual functioning during pregnancy (Daescu, 2023).

#### WEXNER (Stool Incontinence)

WEXNER scores improved modestly between CP1 and CP2 (4.58 → 3.72), suggesting early postpartum recovery in bowel function. No additional late-period data was available for comparison, but the initial improvement coincides with pelvic rehabilitation efforts at one provider, highlighting the importance of linking PROMs to responsive clinical pathways.

#### MIBS (Maternal-Infant Bonding)

Maternal-infant bonding, assessed through the MIBS, improved over time, with scores decreasing from 3.50 to 2.45 between the Early and Late periods and 2.84 to 2.05 across CP3 and CP4 (lower scores indicating better bonding). The improvement suggests that as emotional well-being improves and maternal roles stabilized, bonding with the newborn strengthened. This also reinforces the relationship between psychological well-being and bonding outcomes as seen in previous studies as well (O’Dea GA, 2023).

#### BSES-SF (Breastfeeding Self-Efficacy)

Breastfeeding confidence demonstrated one of the most notable improvements. Between CP3 and CP4, BSES-SF scores increased from 38.98 to 47.81, and over the broader Early to Late comparison, the average rose significantly from 29.38 to 41.77. These gains were likely supported by structured lactation education efforts introduced by multiple providers during the pilot, as well as natural progression in maternal skill and adaptation. The strong upward trajectory highlights both the clinical impact of targeted education and the evolving nature of maternal confidence in the postpartum period, which adds to the existing literature on the relationship between proper lactation education and attitudes towards it (Sandhi A, 2023).

#### Correlations

##### PROM-PROM Correlations

Preliminary analysis of correlations between PROM instruments identified several statistically meaningful associations, as shown in Figure 10:

**Figure 10.**
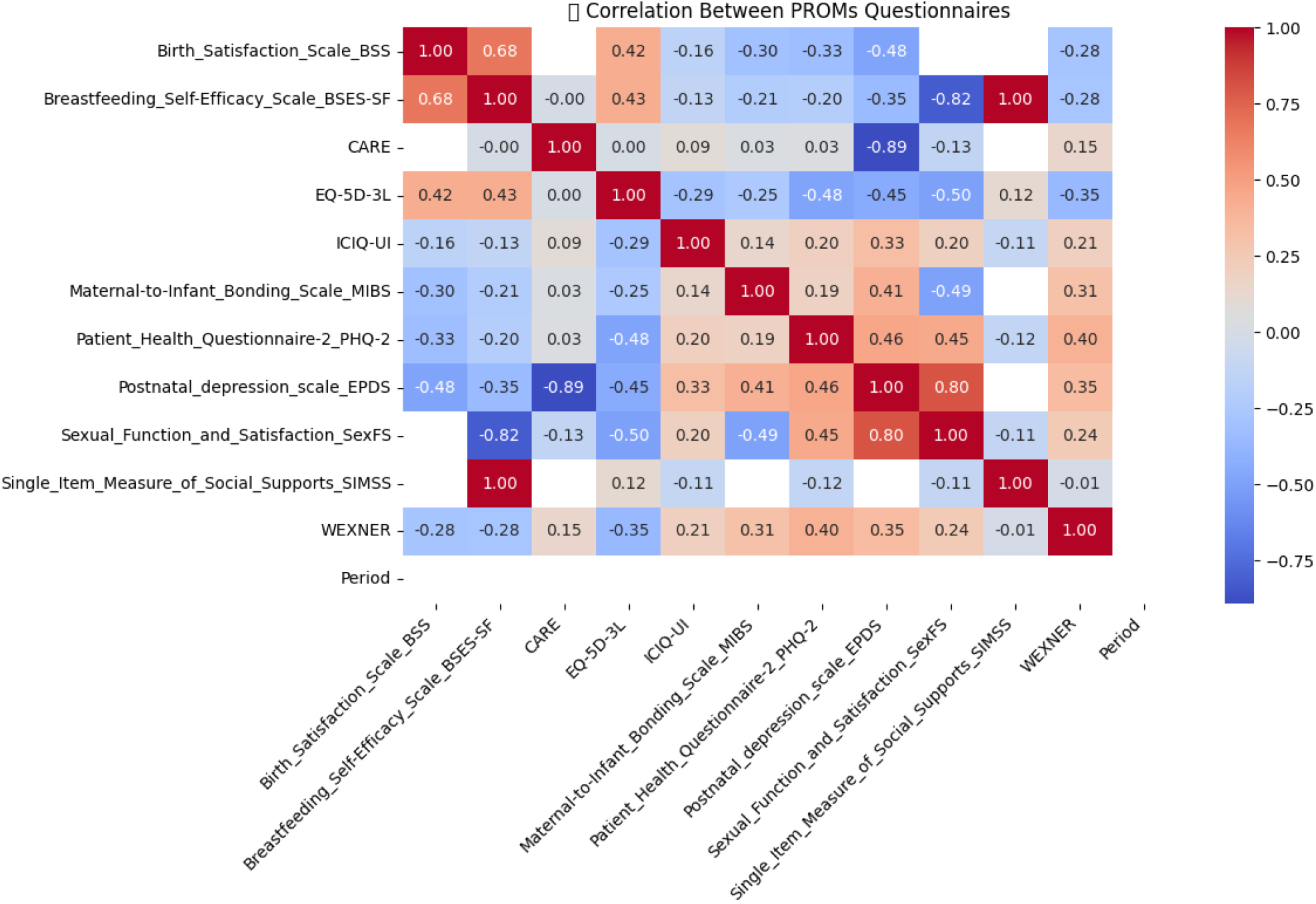
PROM-PROM Correlations

The following are several moderate correlations:

□ Breastfeeding self-efficiency (BSES-SF) correlated positively with Single Item Measure for Social Support (SIMSS) (r=1)
□ Birth Satisfaction Scale (BSS) correlated positively with BSES-SF (r = 0.68) and EQ-5D-3L (r = 0.42), and negatively with EPDS (r = −0.48).
□ EQ-5D-3L demonstrated moderate negative correlations with EPDS (r = −0.45), PHQ-2 (r = −0.48), and SexFS (r = −0.50).
□ PHQ-2 (Depression Screener) was positively correlated with EPDS (r = 0.46), SexFS (r = 0.45), and WEXNER (r = 0.40).
□ MIBS (Maternal-Infant Bonding) showed a moderate positive correlation with EPDS (r = 0.41).

A very strong positive relationship exists between presence of social support and higher levels of breastfeeding confidence. This relationship might be due to how emotional and practical support from family and friends can significantly reduce maternal stress and reinforce positive behaviors, which in turn enhances self-efficacy in breastfeeding. It is worth noting that this correlation was based on a small number of females, and as such may not be as reliable as others. More input is needed to fully corroborate this finding.

The moderate positive correlation between birth satisfaction (BSS) and both breastfeeding confidence (BSES-SF) and general health (EQ-5D-3L) may suggest that women who report feeling well early in their pregnancy—reflected by higher EQ-5D-3L scores—tend to carry this sense of well-being throughout the maternity journey, ultimately culminating in more positive birth experiences and greater postpartum confidence in breastfeeding.

Conversely, the negative correlations between BSS and depression (EPDS) as well as EQ-5D-3L and depression reflect that negative psychological experiences often co-occur with dissatisfaction and reduced quality of life.

The moderate positive correlation between the Maternal-to-Infant Bonding Scale (MIBS) and the Edinburgh Postnatal Depression Scale (EPDS) (r = 0.41) underscores a meaningful relationship between postpartum depression and difficulties in maternal-infant bonding. This finding is consistent with existing literature suggesting that depressive symptoms can interfere with the emotional availability and responsiveness required to establish a healthy bond with the newborn (Saharoy R, 2023).

Taken together, these findings underscore the value of using a multidimensional PROMs framework to capture the complex and interdependent nature of postpartum recovery. By identifying correlations across domains, providers can deliver more personalized care, where elevated risk in one area (e.g., depression) can trigger proactive assessment or intervention in another (e.g., mother-infant bonding). These insights also point to the most impactful areas for intervention, where addressing one domain could improve multiple aspects of well-being. Finally, they support integrating PROMs into clinical dashboards, enabling providers to recognize high-risk profiles and respond holistically.

##### PROM-CROM Correlations

Among the preliminary correlations analyzed between PROMs and CROMs, several strong and statistically significant associations were identified:

□ Maternal-to-Infant Bonding (MIBS) was positively correlated with both failed operative vaginal deliveries (r = 0.40, N = 103) and cases where the fetal head was too large for the maternal pelvis (r = 0.40, N = 103).
□ Breastfeeding Self-Efficacy (BSES-SF) showed a strong inverse correlation with maternal education level (r = −0.37, N = 155).
□ Postnatal Depression (EPDS) was significantly associated with observed signs of maternal infection postpartum (r = 0.35, N = 56).

The PROM–CROM correlation analysis highlights several clinically meaningful associations between patient-reported outcomes and clinician-reported events during the maternity care journey.

The maternal-infant bonding difficulties were linked to failed operative vaginal deliveries and cephalopelvic disproportion—factors commonly associated with birth trauma. These results suggest that obstetric complications may disrupt early bonding experiences and should prompt targeted support for affected mothers. Additionally, difficult births typically result in increased c-section rates and subsequent maternal stress, feelings of loss of control, physical exhaustion, and, at times, neonatal complication (Dekel S, 2019). Further data is required to better understand the psychological impact of c-sections.

The inverse relationship between breastfeeding self-efficacy and maternal education level is particularly interesting. It may reflect higher expectations or greater awareness of breastfeeding challenges among more educated mothers, specifically those who are in the workplace. This finding challenges common assumptions and indicates that confidence-building interventions should not be limited to less educated populations.

In terms of mental health, the positive correlation between postnatal depression and signs of maternal infection reinforces the interplay between physical recovery and emotional well-being. Women who experience postpartum complications may be at elevated risk for psychological distress and may benefit from closer monitoring and mental health screening.

Additional data to be collected at the conclusion of the project are expected to reveal further correlations, particularly regarding the drivers of birth-related CROMs (e.g., cesarean section vs. normal vaginal delivery).

## Conclusion

The Maternity Outcomes Rollout Pilot marks a significant milestone in the adoption of value-based healthcare in Saudi Arabia. Using multiple engagement levers, providers achieved higher female engagement, while sensitive issues, often overlooked in traditional care settings, were more readily surfaced. By embedding PROMs and CROMs into routine care and analyzing the correlations between them, the pilot enabled more personalized care and earlier identification of at-risk females, thereby supporting more timely and targeted interventions.

Importantly, this pilot laid the foundation for broader VBHC initiatives, both in maternity and across other clinical areas. It underscored the need to invest in capabilities, standardize outcomes, and create safe learning environments for providers. As payer-provider collaborations gain momentum in Saudi Arabia, cultural readiness remains crucial. Many providers naturally express concern when engaging with payers, often associating such initiatives with payment pressures. However, this pilot reframed those dynamics by focusing on shared goals—better outcomes, informed choice, and patient-centered care.

Bupa Arabia and Bupa CareConnect is now well-positioned to expand VBHC throughout its provider network. The pilot serves not only as a successful proof of concept but also as a scalable framework for building trust, aligning incentives, and transforming the healthcare landscape for the better.

## Data Availability

All data produced in the present work are contained in the manuscript

